# Epidemiological investigation and prevention control analysis of longitudinal distribution of COVID-19 in Henan province, China

**DOI:** 10.1101/2020.07.25.20161844

**Authors:** Xianguang Yang, Xuelin Chen, Cuihong Ding, Zhibo Bai, Jingyi Zhu, Gege Sun, Guoying Yu

## Abstract

**Objective:** To analyze the vertical distribution of six cities in Henan Province,China from January 21, 2020 to June17, 2020: Xinyang City (including Gushi County), Nanyang City (including Dengzhou City), Zhumadian City (including XincaiCounty), Zhengzhou City (including Gongyi City), Puyang City and Anyang City (including Hua County) corona virus disease 2019(COVID-19) epidemiological characteristics and local prevention and control measures.

**Methods:** Data were collected and analyzed through the COVID-19 information published on the official websites of health commissions of Henan Province and six cities.

**Results:** As of June 17, 2020, the cumulative incidence rate of COVID-19 in Henan province was 1.33/100,000, the cumulative cure rate was 98.27%, the cumulative mortality rate was 1.73%, the age range of diagnosed cases was 5days-85years old, and the male to female ratio was 1.09:1.The confirmed cases of COVID-19 in Henan province were mainly imported cases from Hubei, accounting for 87.74%, of which the highest number was 70.50% in Zhumadian. The contact cases and local cases increased in a fluctuating manner over time.

**Significance:** In this paper, epidemiological characteristics of COVID-19 in Henan province from the outbreak to the effective control within 60 days were analyzed, and effective and distinctive prevention and control measures in various cities were summarized, so as to provide a favorable reference for the further formulation and implementation of epidemic prevention and control and a valuable theoretical basis for effectively avoiding the second outbreak.

**Importance:** Epidemic prevention and control in China has entered the normalized this new stage, this article analyze the epidemiological characteristics of COVID - 19 in henan province, and summarizes the prefecture level and effective disease prevention and control means and measures, the normalized private provide the theoretical reference to the prevention and control work, formulate and carry out, at the same time in other countries and regions of epidemic prevention and control work can also be used for reference.

## 1. Introduction

In December 2019, Wuhan, Hubei province, China reported a number of cases of viral pneumonia of unknown cause[1], whose rapid spread has aroused great concern from the whole society.The virus was confirmed to be a novel coronavirus with a unique genome by sequencing the lower respiratory tract samples collected by the expert group and named as “2019 Novel Corona virus” (2019-nCoV-2),designated by the International Committee for the Classification of Viruses as SARS-CoV-2.The virus novel SARS-CoV-2 spread throughout the world within three months, causing a pandemic and posing a serious threat to human health[2].On February 11, 2020, WHO named the novel coronavirus disease as COVID-19 and declared the infectious disease a public health emergency[3].The COVID-19 is reported to have spread through human-to-human transmission[4].

So far, there is a lack of immunity against the COVID-19 in humans[5].At present, the known source of COVID-19 infection is novel Coronavirus infection (including asymptomatic infection), which is transmitted by respiratory droplets, close contact, aerosol and fecal mouth. Respiratory droplets and close contact are the main transmission routes, and people are generally susceptible.Based on the known epidemiological investigation, the incubation period of this disease is 1-14d, generally 3-7d.Fatigue, dry cough and fever were the main symptoms of common infected persons. A small number of patients were accompanied by nasal congestion, runny nose, sore throat, diarrhea and other symptoms. CT examination showed chest imaging characteristics.Most of the patients with severe illness developed dyspnea or/and hypoxemia 7 days after onset.Critically ill patients may rapidly progress to acute respiratory distress syndrome, septic shock, multiple organ failure and other symptoms.It should be noted that most of the patients with severe and critical diseases had moderate to low fever or no obvious fever symptoms[6-8].

Henan province is adjacent to Hubei province, with Xinyang, Nanyang and Zhumadian as the junction. Zhengzhou, as the capital city of Henan Province, has a large flow of people, so the four cities are COVID-19 high incidence areas in Henan Province.On January 21, 2020 health committee issued reports from Henan province Zhengzhou in Henan province found the first imported COVID - 19 patients, reported on June 26, 24 when Henan cumulative COVID - 19 confirmed cases, 1276 cases, cure the hospital 1254, accumulative total 22 cases died, in addition to March 11, the new case of overseas (Italy) input COVID - 19 confirmed cases, confirmed cases in the province all reset!In this paper, according to the vertical distribution from south to north of Henan province south (Nanyang, Xinyang, Zhumadian), selected (Zhengzhou City), in the north (Anyang, Puyang 6 city COVID - 19 outbreak of epidemiological investigation and analysis of the prevention and control, in order to further to carry out the command of the epidemic prevention and control work, prevent secondary kickback provide theoretical basis for disease[9].

## 2. Materials and methods

### 2.1 Data classification

The data investigated in this paper include age, gender, incubation period, cumulative confirmed cases, cumulative cured and discharged cases, daily new cases/cured cases, number of permanent residents in Henan province and cities, gender and age structure of Henan province and cities, source of confirmed cases and types of confirmed cases.In this paper, the source types of cases were divided into three categories: imported cases (the patient had a history of overseas travel before diagnosis);Contact cases (contact with confirmed or suspected patients before diagnosis);Local case (the patient has not left the city before diagnosis and has no contact history).There are four types of cases: common (mild) cases (mild clinical symptoms, visible imaging/no manifestations of pneumonia);Severe cases (on the basis of the common type, oxygen saturation ≤ 93% under resting state, partial arterial oxygen pressure PaO2/ oxygen absorption concentration FiO2 ≤ 300mmHg, pulmonary imaging showed significant lesion progression within 24-48 hours & GT;50%, etc.);Critical cases (including respiratory failure requiring mechanical ventilation, shock, and other organ failure requiring ICU care);Deaths.The calculated data include: cumulative incidence of COVID-19 (1/100,000) = cumulative confirmed cases of COVID-19 in a city (county included)/permanent resident population in a city by the end of 2018;Cumulative cure rate (%) = cumulative number of cured and discharged COVID-19 cases in a city/cumulative number of confirmed COVID-19 cases in a city;Cumulative mortality rate (%) = The number of COVID-19 deaths in a city/the number of COVID-19 confirmed cases in a city.

### 2.2 Data Sources

According to Chapter third of Henan Statistical Yearbook 2019, the number of permanent residents in Henan province and the 6 cities (including counties) required in this paper is determined.According to the official website of Henan Provincial Health Commission, “The Latest Situation of COVID-19 in Henan Province” column, the overall data of COVID-19 epidemic in Henan province were collected, namely, the number of new confirmed cases/cured and discharged cases, the cumulative number of confirmed cases/cured and discharged cases/death cases, and the number of severe and critical cases.Age, sex, incubation period, source type of COVID-19 cases, number of new confirmed cases, number of cured and discharged cases, and number of deaths in the city were collected through the details of confirmed COVID-19 cases published on the official websites of the municipal Health Commission.

### 2.3 Analytical Methods

Use Excel2016 for data collation and analysis.The maximum value, minimum value and mean value were used to describe the measurement data, and the selection rate and proportion were used to describe the counting data.Statistical charts and tables were used to describe the overall trend of COVID-19 epidemic in Henan province and other cities, as well as the sex and age distribution ratio, case source type and case type composition ratio.

## 3. Results

### 3.1 Basic epidemiological characteristics of COVID-19 in Henan Province

#### 3.1.1 The basic situation of the epidemic in Henan Province

Since the first confirmed imported case was reported in Zhengzhou on January 21, 2020, the daily number of newly confirmed cases in the province has fluctuated with time.On February 4, 2020, the province’s new confirmed cases of peak number of 109 people, then the entire province new daily total number of confirmed cases of fluctuation decline over time, the Henan province on February 24 for the first time, no new confirmed cases, cumulative COVID - 19 confirmed cases, 1272 cases of, March 11, Zhengzhou in case of input outside (Italy).A total of 1,276 confirmed COVID-19 cases have been reported in Henan province as of June 17, 2020, and the distribution of COVID-19 cases in each city is shown in Fig.1.The cumulative incidence was 1.33 per 100,000.

**Figure. 1.**
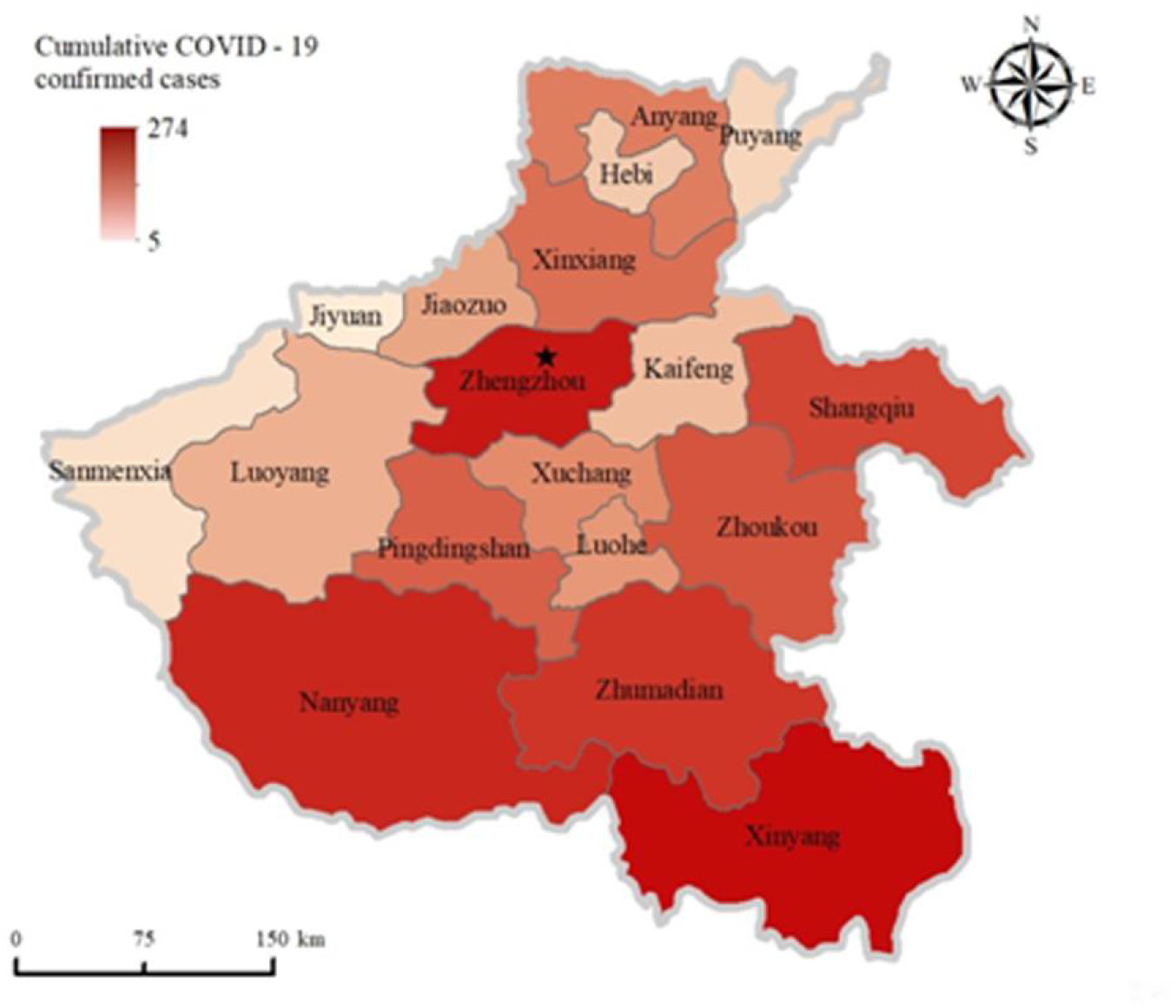
Distribution of COVID-19 in Henan Province, China The figure above indicates the cumulative confirmed COVID-19 cases in Henan Province,China till June 17, 2020. The different colours represent the overall incidence of confirmed COVID-19 cases within the general population within each city of Henan Province, China.The five-pointed star stands for Zhengzhou, the capital of Henan province.

On January 29, 2020, a COVID-19 cured and discharged case was reported for the first time in Henan province. From February 2 to March 8, the total number of COVID-19 cured and discharged cases in Henan province rose with time fluctuation, reaching a peak of 103 cases on February 20.From March 9 to 21, 2020, 1 new COVID-19 patient was cured and discharged from hospital. By March 19, all confirmed COVID-19 hospitalized cases in the province had been cleared to zero, with a total of 1,250 COVID-19 patients cured and discharged from hospital, with a cure rate of 98.19%.

On January 26, 2020, there was one death case of COVID-19 for the first time in Nanyang, Henan province. By March 21, 2020, there had been 22 COVID-19 in Henan province, with a fatality rate of 1.73%.There were 1148 common cases in The province, accounting for 90.25%.There were 64 severe cases, accounting for 5.03%.The total number of critical cases was 38, accounting for 2.99%(Fig.2).

**Figure. 2.**
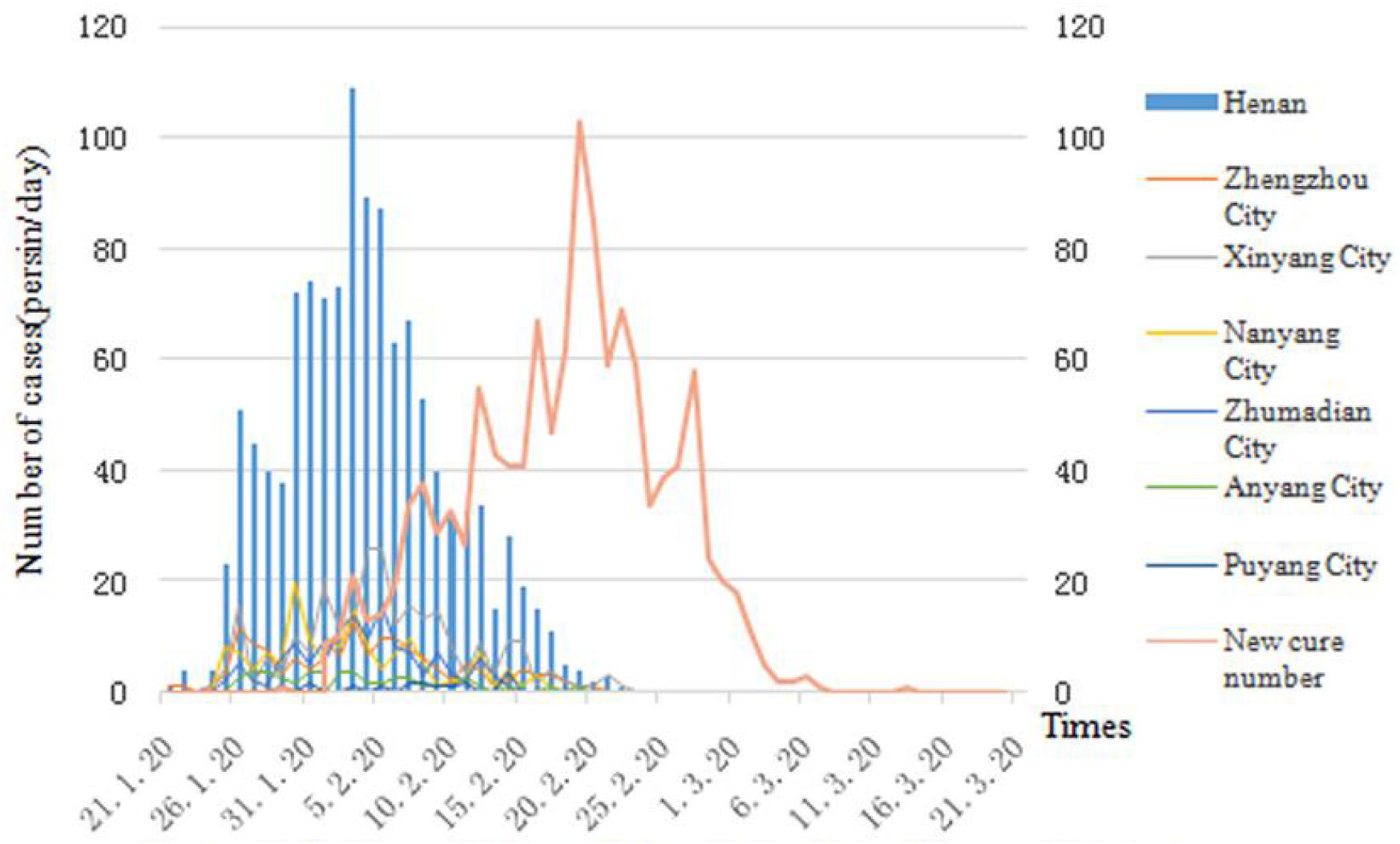
Daily statistics of the epidemic in Henan Province

#### 3.1.2 Basic characteristics of confirmed cases in Henan Province

Due to limited information on confirmed COVID-19 cases published on the official websites of health commissions of various cities, a total of 600 cases in 4 cities (Zhengzhou, Xinyang, Nanyang and Puyang City) with comprehensive information were selected for statistical analysis.Among the 600 confirmed cases, 315 were male and 285 were female, with a sex ratio of 1.11 to 1.The age of confirmed cases ranged from 5 days to 85 years, with the oldest from Zhumadian and the youngest from Xinyang. The average age of confirmed cases was 45 years old, the median age was 46 years old and the majority age was 56 years old.Therefore, it was analyzed that COVID-19 population is generally susceptible to COVID-19, but the middle-aged and elderly population is more likely to be 40-60 years old.Among the confirmed cases of COVID-19, 325 were imported, of which 277 were imported from Hubei, accounting for 85.23%.204 contact cases, accounting for 34%;Local type 73 cases accounted for 12.17%(Fig.3).

**Figure. 3.**
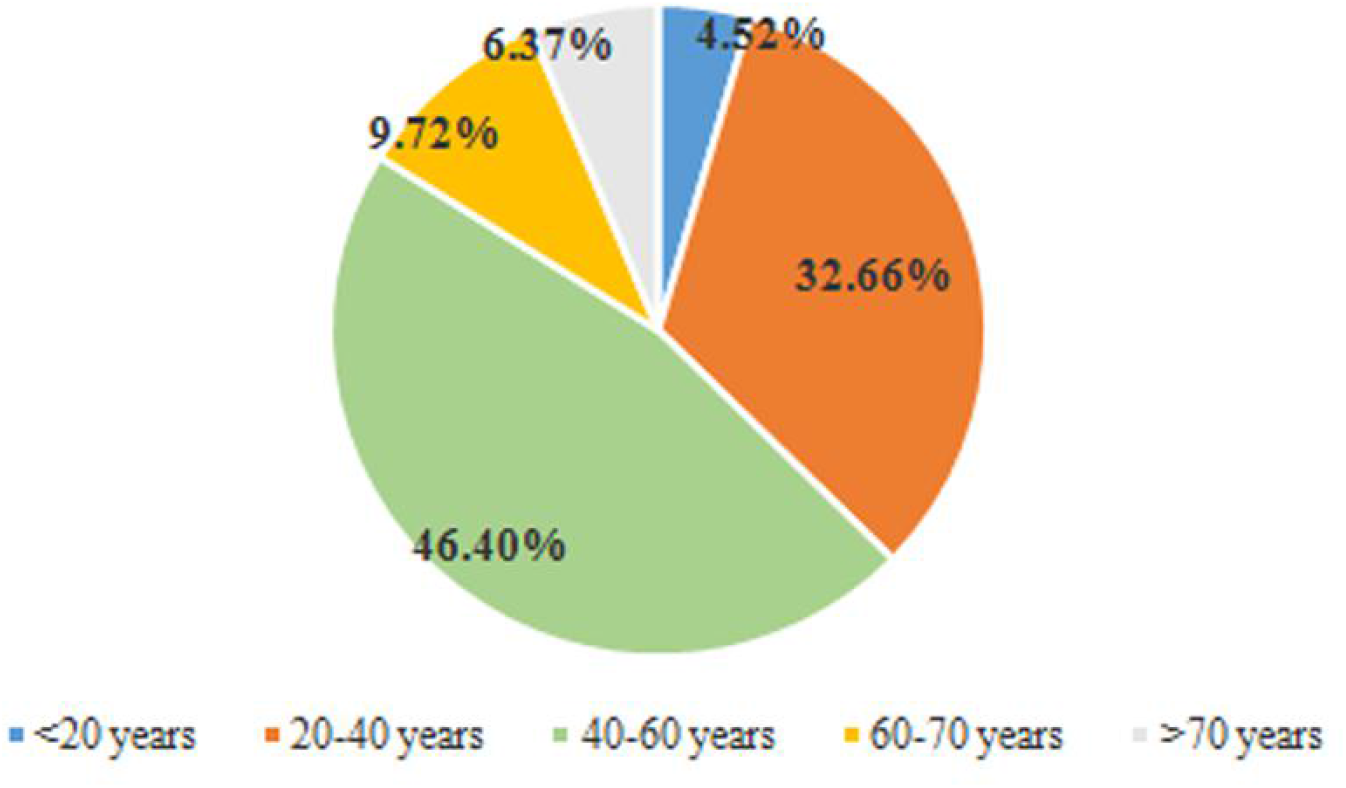
Age ratio of confirmed COVID-19 cases in Henan Province Here is a breakdown of age structure (<60 years old, 20 years old for a period;> 60 years old, 10 years old for a period).

### 3.2 COVID-19 epidemic situation at the municipal level

#### 3.2.1 Variation trend of COVID-19 in each city

As of 24:00 on June 17, 2020, A total of 157 confirmed COVID-19 cases have been reported in Zhengzhou City, with a cumulative incidence of 142/100,000.Xinyang City has reported 273 confirmed COVID-19 cases, with a cumulative incidence of 3.61/100,000.In Nanyang, 154 confirmed COVID-19 cases have been reported, with a cumulative incidence rate of 1.35 per 100,000.In Zhumadian, 139 confirmed COVID-19 cases have been reported, with a cumulative incidence of 175/100,000.Anyang City has reported 52 confirmed cases of COVID-19, with a cumulative incidence of 0.8 per 100,000.Puyang City has reported a total of 17 confirmed COVID-19 cases, with a cumulative incidence of 0.5 per 100,000.Six cities accounted for 62.26% of the province’s total number of confirmed COVID-19 cases. Xinyang had the highest incidence of COVID-19, followed by Zhumadian, Zhengzhou, Nanyang and Anyang City. Puyang City had the lowest incidence of COVID-19.According to the data, the COVID-19 epidemic situation in Henan province as a whole showed a downward trend from north to south based on the analysis of the geographical location and the flow of people in each city.As the capital city of Henan province, Zhengzhou has a complex population and a large number of people moving around. The overall COVID-19 situation is higher than other central regions.On February 23, 2020 first without new Xinyang COVID - 19 confirmed cases, February 24, the province has no new cases confirmed, as of March 10, 2020, the cumulative reported COVID - 19 confirmed cases, 1272 cases of, March 11 people outside input with confirmed cases of Zhengzhou to find one, then as of June 17, 2020, the province had no new confirmed cases, cumulative COVID - 19, 1276 cases of confirmed cases.

#### 3.2.2 Age distribution and sex ratio of confirmed cases

By gender, the male-to-female ratio of confirmed COVID-19 cases was1.38:1 in Xinyang, 0.78:1 in Nanyang, 1.58:1 in Zhumadian, 1.14:1 in Zhengzhou and 0.55:1 in Puyang(Anyang City only disclosed the number of confirmed cases, and did not disclose the detailed information of confirmed cases, so the following contents will not be analyzed in detail).According to the analysis of the sex ratio among cities, the cumulative incidence of COVID-19 among males in Zhengzhou, Xinyang and Zhumadian is higher than that among females. Among them, the cumulative incidence of COVID-19 among males in Xinyang is the highest (4.08 per 100,000).Puyang had the lowest incidence of COVID-19 among males (0.34/100,000).The cumulative incidence of COVID-19 among women in Nanyang and Puyang is higher than that of men, and the highest incidence of COVID-19 among women in Xinyang is 3.11/100,000, while the lowest incidence of COVID-19 among women in Puyang is 1.33/100,000.

According to the age analysis, in order to make a correct and objective analysis of the age composition structure, the age is divided according to the age structure in Henan Statistical Yearbook 2019.The age of confirmed COVID-19 cases ranged from 5 days (Xinyang) to 88 years (Zhumadian), and the age of confirmed COVID-19 cases was concentrated between 15 and 64 years old, accounting for 87.42%. The cumulative incidence of COVID-19 in this age group was the highest (511/100,000) in Xinyang. The cumulative incidence of COVID-19 at age ≤ 14 years was the lowest, with Nanyang having the lowest incidence (0.10/100,000).The results are shown in Table 1.1.The average age of confirmed COVID-19 cases was 42.8 years old in Zhengzhou City, 47.3 years old in Xinyang, 44.4 years old in Nanyang, 43.2 years old in Zhumadian, and 39.5 years old in Puyang.

**Table 1.1.**
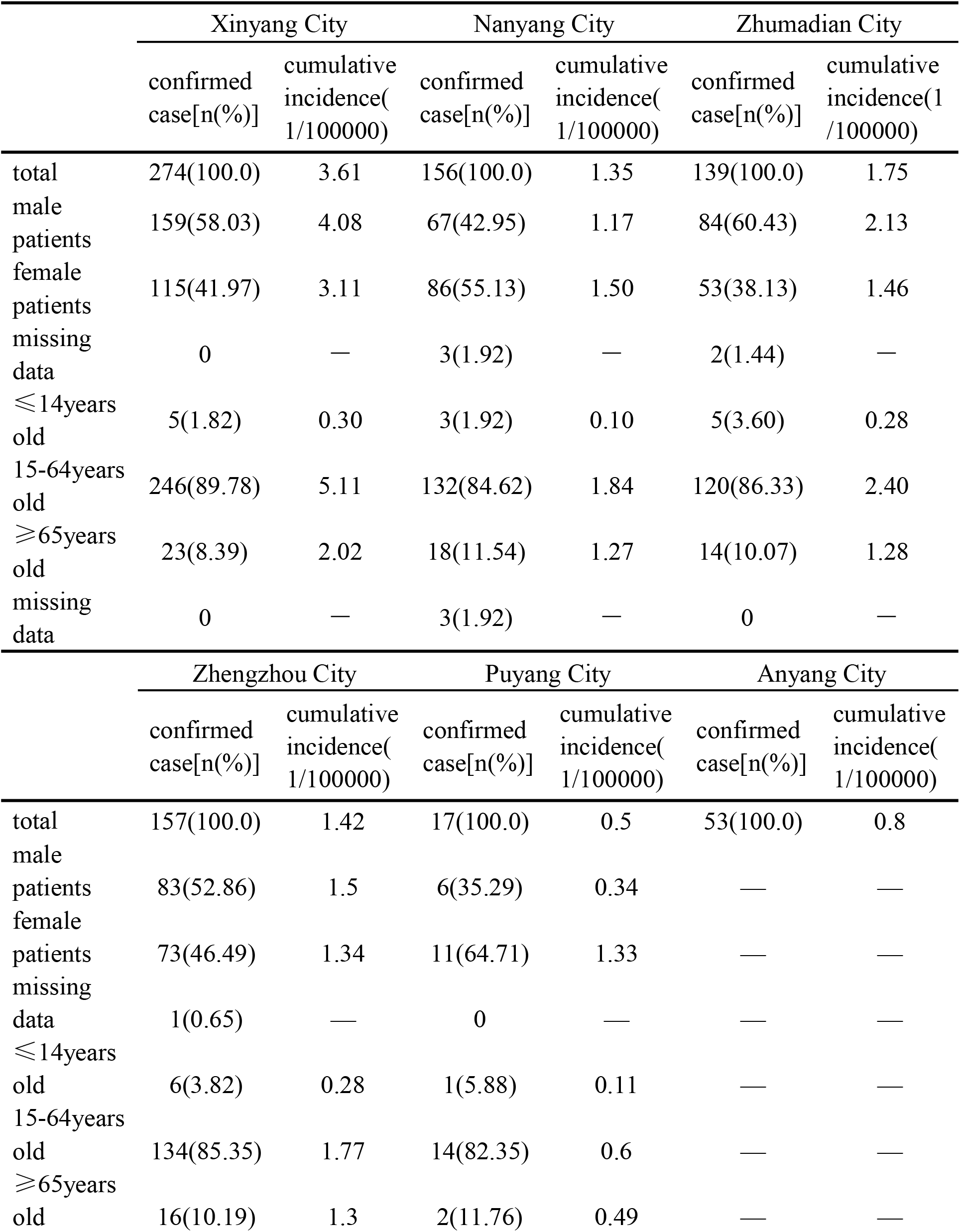
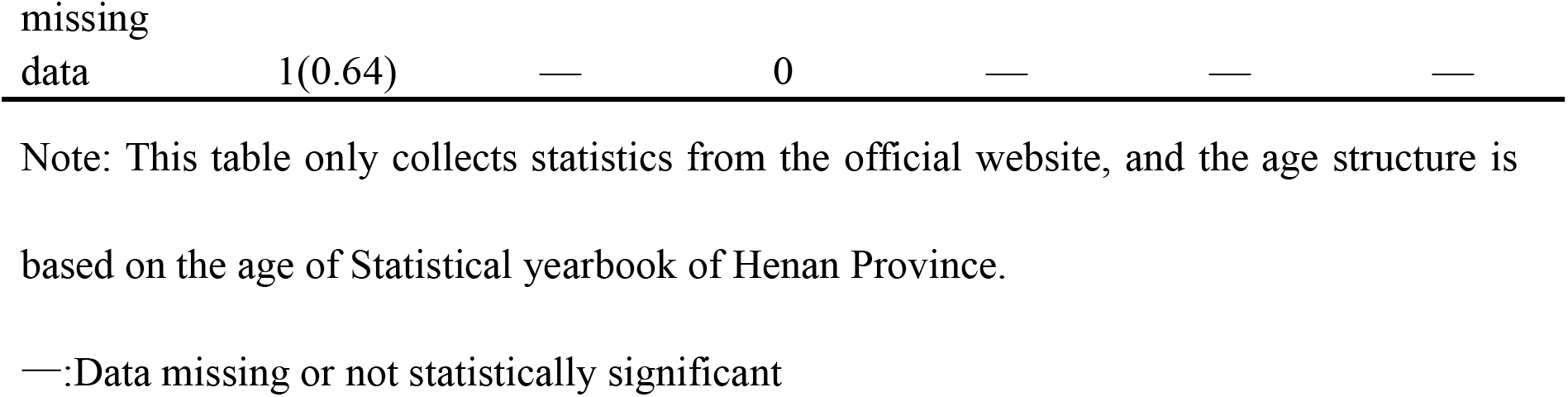
Confirmed cases and cumulative incidence of COVID-19 in six longitudinal cities in Henan province as of June 17, 2020

#### 3.2.3 Source of confirmed cases

As of 24:00 on June 17, 2020, the source type ratio of confirmed COVID-19 cases in each city (input: contact: local) was 4.62:3.79:1 in Xinyang, 5.73:3.60:1 in Nanyang, 8.17:2.42:1 in Zhumadian, 3.96:1.33:1 in Zhengzhou City and 1.5:1.5:1 in Puyang.According to the data, the main sources of confirmed COVID-19 cases are imported cases, among which the highest proportion of imported cases is 70.50% in Zhumadian, followed by Zhengzhou.In Xinyang, imported cases and contact cases accounted for 89.05%, and the ratio of imported cases and contact cases was 1.22:1.The results are shown in Table 1.2.According to the public information of the epidemic notification in Henan Province, confirmed imported cases were selected from Xinyang, Zhengzhou, Zhumadian and Nanyang City for detailed investigation, and the imported cases were subdivided into confirmed imported cases from Hubei and confirmed non-imported cases. The results are shown in Table 2.Hubei accounted for 87.74% of the total imported cases, while Hubei accounted for the majority of COVID-19 imported cases.

**Table 1.2.** Source types of confirmed COVID-19 cases

|  | Xinyang City |  | Nanyang City |  | Zhumadian City |  |
| --- | --- | --- | --- | --- | --- | --- |
|  | confirmed<br>case | percentage<br>[n(%)] | confirmed<br>case | percentage<br>[n(%)] | confirmed<br>case | Percentage<br>[n(%)] |
| imported<br>case | 134 | 49.08 | 85 | 55.19 | 98 | 70.50 |
| contact<br>case | 110 | 40.29 | 53 | 34.42 | 29 | 20.86 |
| local type<br>of case | 29 | 10.62 | 15 | 9.74 | 12 | 8.63 |
| missing<br>data | — | — | 1 | 0.65 | — | — |
|  | Zhengzhou City |  | Puyang City |  | Anyang City |  |
|  | confirmed<br>case | percentage<br>[n(%)] | confirmed<br>case | percentage<br>[n(%)] | confirmed<br>case | percentage<br>[n(%)] |
| imported<br>case | 99 | 63.06 | 6 | 35.29 | — | — |
| contact<br>case | 33 | 21.02 | 7 | 41.18 | — | — |
| local type<br>of case | 25 | 15.92 | 4 | 23.53 | — | — |
| missing<br>data | — | — | — | — | — | — |

Note: This table only collects statistics from the official website, and the age structure is based on the age of Statistical yearbook of Henan Province.
—:Data missing or not statistically significant

**Table 2.**
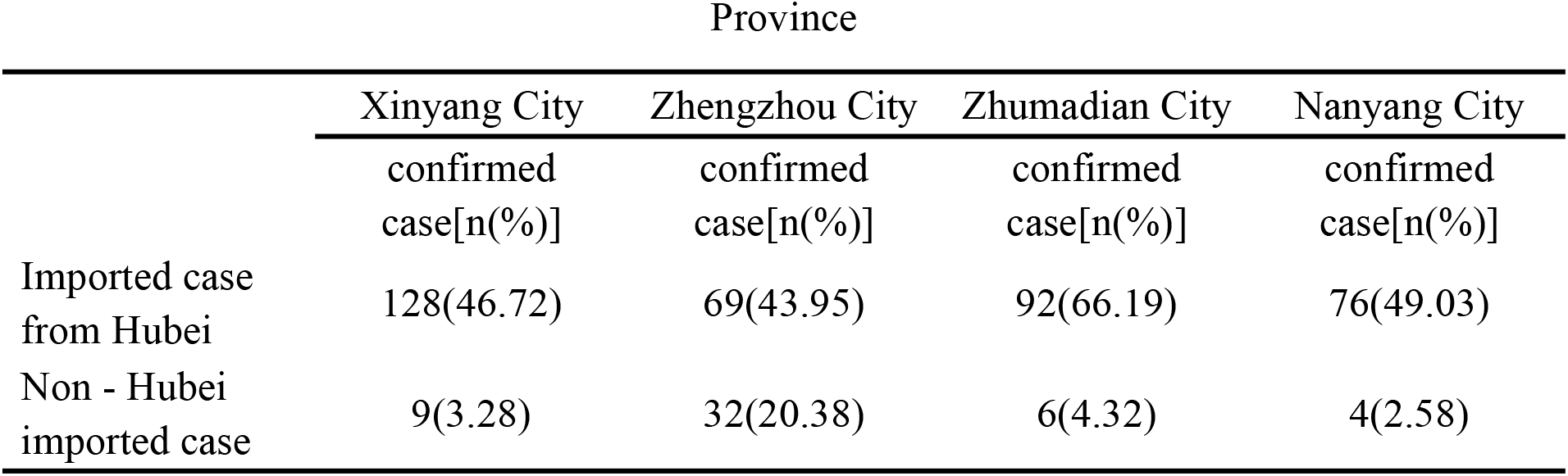
Analysis of imported cases in areas with high epidemic incidence in Henan Province

### 3.3 Epidemic Prevention and control

#### 3.3.1 National epidemic prevention and control

In order to do a good job in the prevention and control of the COVID-19 epidemic in China and safeguard the health and life safety of the people, the National Health Commission has drawn up a novel Coronavirus Pneumonia Prevention and control plan to prevent the spread of the epidemic in light of the epidemic situation and the latest research progress.The health administration departments at all levels are required to coordinate with the CDC, allocate funds and materials for epidemic prevention and control in a timely manner, carry out joint prevention and control work, timely find, timely report, timely feedback and timely analysis.Medical institutions at all levels and of various types are responsible for the detection and reporting of cases, isolation, diagnosis, treatment and clinical management. While preventing and treating cases, they care about people’s mental health problems and do a good job in health education and risk communication with the public.

#### 3.3.2 Main prevention and control measures of Henan Provincial Government

In the face of COVID-19, the Henan Provincial government responded positively, quickly implemented the guidance given in the document of the CPC Central Committee, and added a number of important prevention and control measures with Henan characteristics.First, increase publicity.The government has made the public aware of the seriousness of COVID-19 through TV and radio 24/7, and sent text messages on epidemic prevention and control to the public through mobile phone platforms.Second, strengthen supervision.Since the outbreak of the epidemic, Henan Province has given full play to big data computing.The “Health code” of Henan Province has been recognized by 31 provinces, autonomous regions and municipalities, and 13.98 million people have applied for it. Of citizens to move data through the cloud and the ticket real-name records for screening, grassroots community unit (streets, bring, village, etc.) strict screening area of Hubei province, Xinyang, Nanyang, Zhumadian, Zhengzhou COVID - 19 passengers serious area, take the necessary means to monitor its own home quarantine is greater than 14 d, stop all provincial traffic operations, public supervision and prevention and control of epidemic reporting platform to encourage the populace to supervise to report.Third, fight the epidemic comprehensively.All units at all levels of the health care system will cancel the Spring Festival holiday in 2020, and all staff will arrive at their posts on time to maintain 24-hour communications and respond to the epidemic at any time.130 designated hospitals for MEDICAL treatment of COVID-19 have been identified for timely treatment of COVID-19.

#### 3.3.3 Specific epidemic prevention and control measures

All cities in Henan province promptly launched the first-level response to major health emergencies, and adopted epidemic prevention and control measures with the characteristics of each city.COVID - 19 high incidence area in Henan province (Zhengzhou, Xinyang, Nanyang and Zhumadian City) as an example, the Zhengzhou to open special line 12320 provide consultation service for 24 h free COVID - 19, stop all outside of Zhengzhou passenger service at the same time reduce the city public traffic divisions, each district area residents swept a yard code registration management system, such as have not obey the management focuses on the compulsory isolation.Set up 10 card point in Xinyang, check the traffic of vehicles at the same time for a full sanitizers, temperature detection was carried out on the staff to ensure that no one car, leakage of Hubei direction into the vehicles and personnel are advised to return within the territory of Xinyang, found that patients with fever timely report to the epidemic prevention and control command and arrange to specify the quantities further isolation.All entertainment venues are closed for business. No food in the catering industry, no matter big or small, is allowed to enter public places. Only when there is no abnormality can you enter public places with a mask and a temperature measurement.Nanyang postponed the work of government organs, the resumption of work of various enterprises and the opening of schools, delayed the opening time of administrative examination and approval services under the line, implemented centralized isolation and centralized observation system, and adopted rewarding reporting measures to encourage citizens to supervise surrounding personnel, so as to control the epidemic in an all-round and multi-angle way.Zhumadian and its subsidiaries closed four fire stations, suspended all traffic operations, closed farmers’ markets and entertainment venues.In addition to the above four cities, all other cities have imposed compulsory control on the traffic operation within the city, including subordinate counties, and suspended all kinds of public service places at all levels and all kinds of external purchase and distribution activities, so as to effectively ensure the internal non-proliferation and external import prevention.

## 4. Discussion

COVID-19 is a new and highly infectious disease that will break out in 2020. It has had a great impact on the whole society. It has not only seriously affected people’s life and mental health, but also seriously affected people’s production and life.As a province with a large population and strong personnel mobility, Henan province is close to Hubei Province. As of June 17, 2020, Henan province had a total of 1,273 confirmed cases, with a cumulative incidence rate of 1.32/100,000, higher than that of other provinces and cities outside Hubei, which is closely related to the geographical location of Henan province.Since the first confirmed case was reported in Henan province, the number of new confirmed cases per day in general showed an inverted v-shaped trend, reaching a peak of 109 cases on February 20, and the epidemic reached a stable control after February 24. There was no new case except one imported case from abroad on March 11.Study shows that the population migration and COVID - 19 cumulative disease rate is positively correlated, Zhengzhou as the capital city as the main transport hub city of Henan province, the population of complex personnel fluidity big range, Zhumadian City has 4 place with Wuhan high-speed and next to, Nanyang, Xinyangand Nanyang City in Henan province and Hubei province at the junction, turnover main direction is in Hubei province, due to the convenient transportation only 30 min drive, so both the city personnel will be in Hubei province as the first choice for migrant workers, entertainment, so the above four city of Henan province COVID - 19 outbreaks.Anyang and Puyang City are in the northernmost part of Henan Province, far away from Hubei Province, so the epidemic has been least affected[10].

In terms of the source of confirmed COVID-19 cases, Hubei imported cases dominated the initial stage of the outbreak. Contact cases and local cases fluctuated over time, indicating that the outbreak shifted from external import to internal spread.However, the total proportion of contact cases and local cases was 42.80%, indicating that there was no large-scale spread of the epidemic in Henan province.It is worth noting that one imported case occurred in Zhengzhou on March 11, and the confirmed imported cases from central Hubei accounted for 20.38% of the total cases in Zhengzhou City. Therefore, Zhengzhou City should strengthen the import of other non-Hubei and foreign regions while preventing imported cases from Hubei.

Studies by the COVID-19 Epidemiology Group of the Chinese Center for Disease Control and Prevention showed that the sex ratios of confirmed COVID-19 cases in China, Hubei province and Wuhan were 1.06:1, 1.04:1 and 0.99:1, respectively[11].Zhong Nanshan group of patients in gender studies showed that 41.9% of female patients accounted[12], Xi ‘an, the centers for disease control and prevention reported male patients accounted for 54.3%[13]^[13]^, and Lu Jiahai[14],Kang[15]^[15]^to Guangzhou confirmed cases found women more than men, the prompt COVID - 19 men and women are prone to infection in the gender, but confirmed cases of gender distribution is different in different areas.For example, male patients in Zhengzhou City, Xinyang and Zhumadian City were more than female patients, while female patients in Puyangand Nanyang City were more than male patients.

Since January 21, the province has reported 1,273 confirmed COVID-19 cases (including one imported from abroad) and 22 deaths. On March 15, as the last COVID-19 patient in Xinyang was discharged from hospital on March 14, all local hospitalized patients in the province have been cured and discharged, realizing zero cases!At present, the COVID-19 epidemic in Our province has been effectively controlled, but do not relax your vigilance!With the return of overseas students and the return of overseas staff, while preventing the spread of asymptomatic infected persons in Our province, we should strictly control the import of external cases, strictly control the risk of imported cases, strengthen the epidemic prevention and control work in our province, and earnestly achieve the goal of”internal anti-diffusion, external anti-input”.

## Data Availability

All data generated or analyzed during this study are included in this article.

## Acknowledgment

We thank Dr. Honglei Zhu for his guidance on figure.1 in the manuscript.

## References

[1] Commission WMH. Report on current pneumonia epidemic situation in the city (in Chinese). 2020.http://wjw.wuhan.gov.cn/front/web/showDetail/2019123108989 (accessed March 2, 2020).

[2] Rothan HA, Byrareddy SN. The epidemiology and pathogenesis of coronavirus disease (COVID-19) outbreak. J Autoimmun 2020; 109:102433.

[3] WHO. Novel Coronavirus (2019-ncov) - Situation Report – 22, 2020. https://www.who.int/docs/default-source/coronaviruse/situation-reports/20200211-sitrep-22-ncov.pdf?sfvrsn¼fb6d49b1_2. (accessed March 2, 2020).

[4] Li Q, Guan X, Wu P, Wang X, Zhou L, Tong Y, et al. Early Transmission Dynamics in Wuhan, China, of Novel Coronavirus-Infected Pneumonia. N Engl J Med 2020; 382:1199–207.

[5] AH A, F AO. Novel SARS-CoV-2 outbreak and COVID19 disease; a systemic review on the global pandemic. Genes & Diseases 2020.

[6] Huang C, Wang Y, Li X, Ren L, Zhao J, Hu Y, et al. Clinical features of patients infected with 2019 novel coronavirus in Wuhan, China. Lancet (London, England) 2020; 395.

[7] Riou J, Althaus CL. Pattern of early human-to-human transmission of Wuhan 2019 novel coronavirus (2019-nCoV), December 2019 to January 2020. Euro Surveill 2020; 25.

[8] China NHCO. Notification on the issuance of the COVID-19 Protocol (trial seventh edition). 2020.

[9] Province HCOH. Update on COVID-19 in Henan province as of 24:00, March 21. 2020.

[10] Baidu migration: Destinations moved out of Hubei Prov-ince. 2020.

[11] CDC. CERM. Epidemiological characteristics of COVID - 19. Chinese Journal of Epidemiology 2020.

[12] Guan W, Ni Z, Hu Y. Clinical characteristics of 2019 novel coronavirus infection in China. 2020.

[13] Yao B, Kun L, Zhijun C, Baozhong C, Zhongjun S. Early transmission dynamics of COVID-19 in Shanxi Province. Chinese Journal of Nosocomiology 2020; 30:834–8.

[14] Xuanzhuo W, Conghui L, Zhihui L, Huan H, Xiaomin C, Qianlin L, et al. Preliminary analysis of the early epidemic and temporal and spatial distribution of COVID-19 in Guangdong Province. Journal of Tropical Medicine 2020; 20:427–30.

[15] M K, J W, W M. Coronavirus - COVID-19; Evidence and characteristics of human-to-human transmission of SARS-CoV-2 (Updated February 17, 2020). Medical Letter on the CDC & FDA 2020.

